# Ultrasound Guided Brachial Plexus Block in Upper Extremity Surgeries: Experiences from a Tertiary Care Hospital of Bangladesh

**DOI:** 10.1101/2021.07.04.21259905

**Authors:** Mohammad Ali, Fatema Johora, Sabiha Mahboob, Sonia Nilufar, Mst. Arifa Shirin

## Abstract

Brachial plexus block has been gained popularity after its introduction because of provision of quality anesthesia and optimal postoperative analgesia in upper arm surgeries, and introduction of ultrasound guidance in recent years further enhances the practice because of efficacy and safety of procedure. This retrospective observational study evaluated the outcome and safety of ultrasound-guided brachial plexus block (supraclavicular and interscalene approach) in a tertiary care hospital of Bangladesh over 3-years period (January 2017 to December 2019) through analyzing preoperative anesthesia evaluation form, anesthesia documents and postoperative records. 113 patients were covered during the study period, of which 59 (52.2%) were males and 54 (47.8%) were females. Mean age of the patients were 38.94 ± 20.37 years. Majority of the patients (63.7%) were in ASA grading I. Operative time for surgeries were 115.30 ± 35.19 minutes. duration of sensory and motor block were 618 ± 55 minutes and 450 ± 74 minutes respectively. And first dose of postoperative analgesic was given after 705 ± 94 minutes of surgery. Tachycardia and hypertension were observed in 4.4% patients, where failure of motor blockade and puncture of subclavian vein were recoreded 1.8% and 0.9% respectively. Current study found that with ultrasound guidance in skilled hands, brachial plexus blockade through combination of supraclavicular and interscalene approaches can provide quality anesthesia and analgesia with minimal complications.

## Introduction

Brachial plexus block was first performed under direct visualisation after surgical exposure in 1884.^1^ Over the ensuing years, a variety of techniques, modifications, and advancements have made brachial plexus block one of the regional anesthetic techniques most frequently used in contemporary practice of surgical anesthesia and postoperative pain management in upper arm surgeries because of avoidance of side effects of general anesthesia, optimal postoperative analgesia, less requirement of opioid analgesic, less duration of hospital stay and cost-effectiveness. It can be extremely applicable in patients with significant co-morbidities e.g. significant cardiopulmonary dysfunction, morbidly obese and those with potential difficulties.^2, 3^ Successful brachial plexus blocks rely on proper techniques of nerve localization, needle placement, and local anesthetic injection.^4^ Supraclavicular, infraclavicular, axillary and interscalene approaches are used for brachial plexus block and the very definition of success varies widely. With each anatomic approach to the brachial plexus, several methods of needle localization have been described for injecting after fascial clicks, mechanical paresthesia, electrical stimulation, transarterial injection, fanning injections, use of catheters, and using various imaging modalities like fluoroscopy and ultrasound.^5^ Failure of the procedure, nerve injury and intravascular injection are the most common complications.^6, 7^ In recent years, the use of ultrasound brachial plexus block has been widely used and considered as technique providing best quality of regional anesthesia, irrespective of the approach. Visualization of the target structures (nerve, sheath, interfascial space) as well as the visualization of the needle and the spread of local anesthetic after injection in ultrasound guided technique has increased the success rate of the procedure and reduced risk of complications, block performance time, number of needle insertions and volume of local anesthetic agents.^8, 9, 10^

This current study aims to share the experiences of ultrasound-guided brachial plexus block in a tertiary care hospital of Bangladesh over 3-year period in order to provide the insight of outcome and safety of the procedure.

## Material &Methods

### Place and duration of the study

This was a retrospective observational study, conducted in a tertiary level hospital of Dhaka city from January 2017 to December 2019.

### Procedure

This study was carried out by retrospectively analyzing data of patients, who underwent upper extremity surgery using brachial plexus block (supreclavicular and interscalene approach) in Asgar Ali Hospital Ltd between January 2017 to December 2019. Demographic data, such as age, gender, ASA (American Society of Anesthesiologists) grading, name of operations, diagnoses of patients, were recorded from the preoperative anesthesia evaluation form. Patients having ASA grading III or IV were excluded from thestudy. Data of total 113 patients were analyzed.

In Asgar Ali Hospital, ultrasound-guided brachial plexus applications were performed routinely in the operation room. Standard monitoring was done preoperatively through blood pressure, heart rate, ECG and peripheral oxygen saturation (SpO2) measurements. All the procedures were carried out with portable ultrasound machine in musculoskeletal mode. 22 G nerve block needle was used for the procedure. Procedure and operation times were examined through anesthesia documents retrospectively and recorded as study data. The operation time was defined as the time from skin incision to the last suturing.

Anesthesia documents and postoperative records were examined retrospectively, and applied block type (interscalene and supraclavicular block), local anesthetics and adjuvants used for the block, motor and sensorial block times, timing of first dose of postoperative analgesic were recorded as study data. Patients were briefed about the procedure and written informed consent was taken. Intravenous access was established and suitable IV fluid was started. With monitoring of pulse oxymetry, blood pressure and ECG, patients were positioned to get a proper access to the side of neck and supraclavicular area. With all aseptic precautions, brachial plexus was visualized with help of ultrasound in both supraclavicular and interscalene route. 0.25% bupivacaine and 1% lidocaine with adrenaline were the local anesthetic to be used. Doses of bupivacaine never exceeded 2mg/kg and for lidocaine with adrenaline less than 5 mg/kg was used. 0.1mg/ kg of dexamethasone was used as adjuvant. A total volume of 40 ml was made with normal saline added to drugs mentioned. With the help of ultrasound, 30 ml drugs was injected in supraclavicular route and 10 ml was injected in interscalene route. Both routes were used for reducing the incidence and intensity of tourniquet pain. Onset of block was checked by taking history of tingling sensation, hot and cold sensation and examination for sensory and motor block were done. Motor block time is defined as the time from the time when the Modified Bromage Scale (0 = no motor block, 1 = no shoulder abduction but elbow flexion was present, 2 = both shoulder abduction and elbow flexion are absent, 3 = full motor block) is 1 and above until the motor block disappears completely. The sensorial block time is defined as the time between the disappearance of the sensation of pain and the reappearance of the sensation of pain in one of the dermatomes with a pinprick test performed on one of the C4-T1 dermatomes. Patient was monitored for any complication. Complications, such as bradycardia, tachycardia, hypotension, hypertension, pain sensation during the operation, poor motor block were examined retrospectively through anesthesia documents and recorded as study data. There was no incidence of ipsilateral diaphragmatic paresis, conjunctival congestion, Horner’s syndrome, pneumothorax, hemothorax, laryngeal and phrenic nerve block. Complications observed during the period until the motor and sensory block disappeared postoperatively were examined retrospectively through anesthesia documents and postoperative records, and recorded as study data.

### Statistical analysis

Data was compiled, presented and results are expressed as mean ± SD and percentage.

## Result

One hundred and thirteen patients were covered during the study period, of which 59 (52.2%) were males and 54 (47.8%) were females. Mean age of the patients were 38.94 ± 20.37 years. Majority of the patients (63.7%) were in ASA grading I.

Table II showed that Operative time for surgeries were 115.30 ± 35.19 minutes, duration of sensory and motor block were 618 ± 55 minutes and 450 ± 74 minutes respectively. And first dose of postoperative analgesic was given after 705 ± 94 minutes of surgery.

**Table I:** Demographic data.

|  |  |  |
| --- | --- | --- |
| **Age (year)** | **(Mean ± SD)** |  |
| | $38.94 \pm 20.37$ | |
| **Gender** | **Frequency** | **Percentage** |
| Male | 59 | 52.2% |
| Female | 54 | 47.8% |
| **ASA grading** | **Frequency** | **Percentage** |
| I | 72 | 63.7% |
| II | 41 | 36.3% |
| III | 00 | 0.0% |
| IV | 00 | 0.0% |

**Table II:** Effect of characteristics of brachial plexus blockade (N=113)

|  |  |
| --- | --- |
| **Operative time (minute)** | **(mean ± SD)** |
| | $115.30 \pm 35.19$ |
| Onset (minutes) | $10.06 \pm 1.53$ |
| Duration (minutes) | $618 \pm 55$ |
| **Complete Motor block** |  |
| Onset (minutes) | $17.7 \pm 2$ |
| Duration (minutes) | $450 \pm 74$ |
| **First analgesic given (minutes)** | $705 \pm 94$ |

Tachycardia and hypertension were observed in 4.4% patients, where failure of motor blockade and puncture of subclavian vein were 1.8% and 0.9% respectively (Table III).

**Table III:**
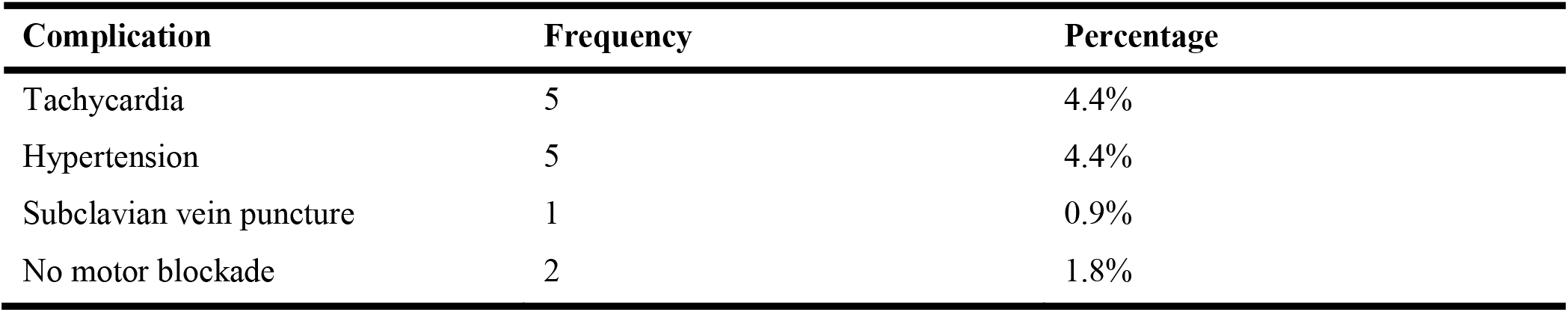
Incidence of complications (N=113)

## Discussion

Brachial plexus block remains the only practical alternative to general anesthesia for significant surgery on the upper limb. Provision of superior quality anesthesia and analgesia while avoiding airway instrumentation, hemodynamic consequences of general anesthesia, favorable postoperative recovery profile along with patient’s satisfaction and cost-effectiveness are the factors for growing popularity of brachial plexus block for upper extremity surgeries. The choice of technique depends on the type of surgery, experience of the anesthesiologist, perceived complications of the individual block, and the patient’s well being.^11,12^ Although ultrasound as a nerve localization technique has a better safety profile and several other advantages over other ‘conventional’ techniques, serious complications are not completely avoided.^12^ In Bangladesh, fewer studies were conducted in this regard, and current study was designed to explore efficacy and safety of ultrasound guided brachial plexus block in upper arm surgeries in a tertiary care hospital of private setting.

The supraclavicular block is ideal in providing a dense, rapid onset, and efficient anaesthesia and analgesia for procedures from mid-humerus proximally, to those performed on the hand distally. And has the most widespread extent of sensory blockade among all the brachial plexus approaches. Interscalene block provides reliable anaesthesia and analgesia for procedures (open and arthroscopic) involving the shoulder joint, lateral two-thirds of the clavicle and proximal humerus. It effectively blocks the proximal nerve roots, distal cervical plexus (supraclavicular nerves), and important nerves such as the suprascapular, which exit proximally from the plexus.^13^ In current study, combination of supraclavicular and interscalane approaches was used. Shorter onset of action and longer duration of sensory and motor block were cited as advantages of ultrasound-guided brachial plexus blockade.^12^ Onset of sensory block was 10.06 ± 1.53 minutes and that was concordance with relevant literatures.^14, 15^ Duration of sensory block was 618 ± 55 minutes and it was similar to observation of Altinay et al.^16^ Onset of motor block was 17.7 ± 2 minutes which was slower than previous studies^15, 16^ probably less drug concentration was used in current study to avoid local anesthesia toxicity. Duration of motor block was 450± 74 minutes and that was comparable to previous literature.^16^

Orthopedic upper extremity surgeries are considered as having a high incidence of postoperative pain.^17^ One of the important advantages of brachial plexus block over general anesthesia for upper extremity surgery is the long duration of postoperative analgesia.^18^ In current study, along with anesthetic agents bupivacaine and lidocaine, adrenaline and dexamethasone were used. Adrenaline delays absorption of local anesthetic through vasoconstriction as well as works directly on spinal cord for pain suppression through α_2_ adrenoceptor stimulation.^19, 20^ Addition of long acting glucocorticoid, dexamethasone potentiates analgesia by both local and systemic effects on nerve fibers.^21, 22^ Duration of postoperative analgesia was 705 ± 94 minutes and that was similar to related study^18^

The vast majority of orthopedic, plastic and vascular surgeries can be performed safely using supraclavicular and interscalene block. These blocks provide efficacy with fewest significant complications including reduced incidence and intensity of tourniquet pain and their best performance can be achieved through ultrasound.^11, 13^ Tachycardia and hypertension were found as common complications in current research due to effects of adrenaline on heart and blood vessels.^20^ It would be better to avoid adrenaline in hypertensive patients in future. The improved safety offered by ultrasound in location and in particular the visualization of pleura and needle position has been the turning point in the increased use of this approach. Although the incidence of pneumothorax, phrenic nerve block and puncture of subclavian vessels are reduced but reports still found in other study.^12^ In this study, puncture of subclavian vein was happened in one case which was managed successfully and there was no incidence of subsequent hemothorax. Rate of failure of motor block was 1.8% in this research whether it was found 13-20% in comparable works.^15, 24^ Actually the success of ultrasound-guided regional technique largely depends on skill and experience of anesthesiologist.^25, 26^

## Conclusion

Brachial plexus block is an excellent alternative of general anesthesia for a wide variety of upper limb surgeries as well as optimal postoperative analgesia with minimal complications and increased patient satisfaction. Ultrasound guidance with real-time needle visualization in relation to anatomic structures and target nerves makes brachial plexus safer and more successful. Current study showed with ultrasound guidance in skilled hands, brachial plexus blockade through combination of supraclavicular and interscalane approaches, can provide quality anesthesia and analgesia with minimal complications. By combining ease of performance with efficacy and safety, it is hoped that anesthesiologists will consider in adopting this technique in their day to day practice.

## Data Availability

All data are available on request

